# CoViD–19: Meta-heuristic optimization based forecast method on time dependent bootstrapped data

**DOI:** 10.1101/2020.04.02.20050153

**Authors:** Livio Fenga, Carlo Del Castello

## Abstract

A compounded method – exploiting the searching capabilities of an operation research algorithm and the power of bootstrap techniques – is presented. The resulting algorithm has been successfully tested to predict the turning point reached by the epidemic curve followed by the CoViD–19 virus in Italy. Futures lines of research, which include the generalization of the method to a broad set of distribution, will be finally given.

## 1. Introduction

In general, predicting the time of a peak conditional to a set of time dependent data is a non trivial task. Often carried out in a multi-tasking fashion, requiring the availability of time and resources, the correct estimation of future turning points can be important in many instances but becomes crucial in the case of epidemic events. These are the typical circumstances when the forecasting exercise is conducted on-line and on a time series exhibiting a small sample size. However, under these conditions, the problem might become particularly complicated since statistical methods usually employed for this purposes – for example of the type hidden Markov (see, e.g. Hamilton (1989) and Koskinen and Ö ller (2004)) or non parametric (see, e.g., Delgado and Hidalgo (2000)) models – not only are very demanding in terms of building and tuning procedures, but typically require the availability of a “long” stretch of data. In addition to that, the time series related to epidemics usually show highly non-linear dynamics, which, if not pre-processed, make them not suitable for standard linear models. On the other hand, attempting to fit non-linear models – e.g. of the type Self Exciting Threshold Autoregressive (see, for example Clements et al. (2003)) or artificial neural network (Hassoun et al. (1995)) – it is not a viable options, due to the above mentioned sample size issues. In any case, when an ill-tuned model is fitted on a time series, reliable outcomes should not be reasonably expected. Therefore, an approach able to perform under the above outlined conditions, is proposed. In essence, the problem is solved by building a unified framework in which two powerful techniques – belonging to two different branches of computational statistics – are sequentially employed to lower the amount of uncertainty embedded in the observed data and to find a (possibly global) optimum through which the “best” statistical distribution for the data set at hand is found.

## 2. The unified framework

As above stated, the approach studied in this paper, is rooted in a unified framework in which two powerful paradigms are exploited. The first one, which belongs to the so called computer intensive statistical methods, is the bootstrap, which will be detailed in 4. By using this technique, an high number of “bona fide” replications of the original data are generated. In essence, each of the bootstrap series obtained “mimics” the observations recorded, so that the number of series observed – which *in real* life is typically equal to one – becomes (very) high. Repeating a mathematical operation (e.g. the computation of an estimator) B times makes possible *i*) the assessment of the degree of uncertainty associated with the obtained estimations and *ii*) less biased estimators. The latter goal is achievable by design of the bootstrap method, as through its replications the use of central tendency functions, such as mean or median, are possible. The second tool employed, is an optimization method for the selection of the “best” parametrization of a class of statistical distribution commonly used in the literature. In practice, this step is performed in the so called bootstrap world, meaning that it is sequentially repeated for each bootstrap sample. By doing so, the degree of uncertainty associated with the selected distribution is lower than the one obtainable by processing just one set of data (the real observations).

## 3. Data and contagion indicator

This paper uses the official data diffused daily by the National Institute of Health – an agency of the Italian Ministry of health – and the Department of Civil Protection. The data set includes 38 daily data points collected at national level during the period starting from January 19^*th*^ to March 27^*th*^. The used indicator – which will refer to as the variable of interest – is obtained by subtracting, for each day, from the total number of people tested positive of Corona virus the number of the deaths and of the recovery.

## 4. The Resampling Method

The choice of the most appropriate resampling method is far from being an easy task, especially when the identical and independent distribution *iid* assumption (Efron’s initial bootstrap method) is violated. Under dependence structures embedded in the data, simple sampling with replacement has been proved – see, for example Carlstein et al. (1986) – to yield suboptimal results. As a matter of fact, *iid*–based bootstrap schemes are not designed to capture, and therefore replicate, dependence structures. This is especially true under the actual conditions (small sample sizes and strong non-linearity). In such cases, selecting the “right” resampling scheme becomes a particularly challenging task as many resamplig schemes are not designed to capture the dynamics typically found in epidemiology. As an example, the well known resampling method called sieve bootstrap – introduced by Bühlmann et al. (1997) – cannot be employed due to the quadratic shape almost always found in this type of time series.

In more details, while in the classic bootstrap an ensemble **Ω** represents the population of reference the observed time series is drawn from, in *MEB* a large number of ensembles (subsets), say {***ω***_1_, …, ***ω***_*N*_} becomes the elements belonging to **Ω**, each of them containing a large number of replicates {*x*_1_, …, *x*_*J*_}. Perhaps, the most important characteristic of the *MEB* algorithm is that its design guarantees the inference process to satisfy the ergodic theorem. Formally, denoting by the symbol | ***·*** | the cardinality function (counting function) of a given ensemble of time series {*x*_*t*_ *∈* ***ω***_*i*_; *i* = 1, …, *N*}, the *MEB* procedure generates a set of disjoint subsets **Ω**_***N***_ *= ω*_1_ *∩ ω*_1_ *· · · ∩ ω*_*N*_ s.t. 𝔼**Ω**_***N***_ *≈ µ*(*x*_*t*_), being *µ*(*·*) the sample mean. Furthermore, basic shape and probabilistic structure (dependency) is guaranteed to be retained 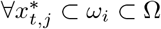.

*MEB* resampling scheme has not negligible advantages over many of the available bootstrap methods: it does not require complicated tune up procedures (unavoidable, for example, in the case of resampling methods of the type Block Bootstrap) and it is effective under non-stationarity. *MEB* method relies on the entropy theory and the related concept of (un)informativeness of a system. In particular, the Maximum Entropy of a given density *δ*(*x*), is chosen so that the expectation of the Shannon Information *H* = 𝔼 (*−* log *δ*(*x*)), is maximized, i.e.

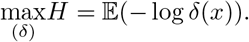

Under mass and mean preserving constraints, this resampling scheme generates an ensemble of time series from a density function satisfying (4). Technically, *MEB* algorithm can be broken down in the 8 steps below detailed.

1. A sorting matrix of dimension *T ×* 2, say *S*_1_, accommodates in its first column the time series of interest *x*_*t*_ and an Index Set – i.e. *I*_*ind*_ = {2, 3, …, *T*} – in the other one;
2. *S*_1_ is sorted according to the numbers placed in the first column. As a result, the order statistics ***x***_**(*t*)**_ and the vector *I*_*ord*_ of sorted *I*_*ind*_ are generated and respectively placed in the first and second column;
3. compute “intermediate points”, averaging over successive order statistics, 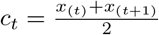, *t* = 1, … *T −*1 and define intervals *I*_*t*_ constructed on *c*_*t*_ and *r*_*t*_, using *ad hoc* weights obtained by solving the following set of equations: i)

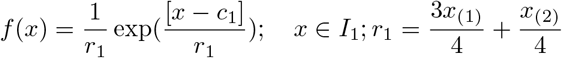 ii)

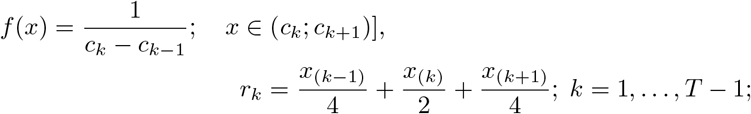 iii)

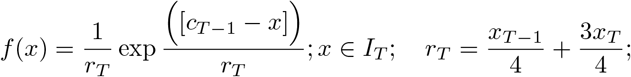
4. from a uniform distribution in [0, 1], generate *T* pseudorandom numbers and define the interval *R*_*t*_ = (*t/T*; *t* + 1*/T*] for *t* = 0, 1, …, *T −* 1, in which each *p*_*j*_ falls;
5. create a matching between *R*_*t*_ and *I*_*t*_ according to the following equations:

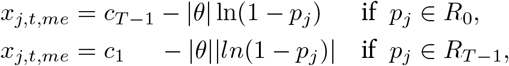

so that a set of *T* values {*x*_*j,t*_}, as the *j*^*th*^ resample is obtained. Here *θ* is the mean of the standard exponential distribution;
6. a new *T ×* 2 sorting matrix 𝒮_2_ is defined and the *T* members of the set {*x*_*j,t*_} for the *j*^*th*^ resample obtained in Step 5 is reordered in an increasing order of magnitude and placed in column 1. The sorted *I*_*ord*_ values (Step 2) are placed in column 2 of 𝒮_2_;
7. matrix *S*_2_ is sorted according to the second column so that the order {1, 2, …, *T*} is there restored. The jointly sorted elements of column 1 is denoted by {*x*_*S,j,t*_}, where *S* recalls the sorting step;
8. Repeat Steps 1 to 7 a large number of times.

## 5. Bootstrap driven forecast optimization

This section aims to define an alternative method to forecast our variable of interest using an optimization approach to fit a set of distribution functions on bootstrap replications.

The variable of interest is assumed to approximately describe a logistic function, scaled by a normalizing parameter *h* (representing the asymptotic number of cases) and its derivative is a Gaussian function re-scaled by the parameter *h*.

**Fig. 1.**
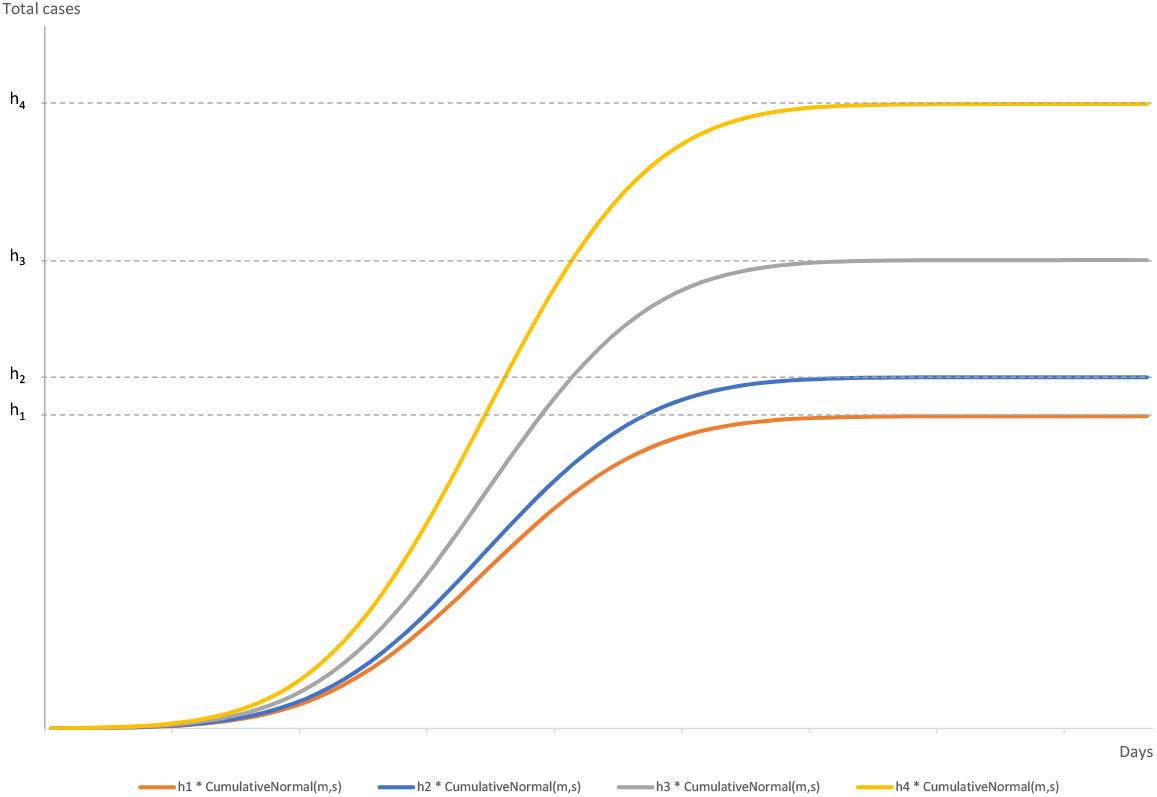
COVID19 Active cases

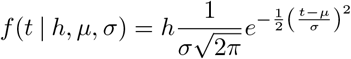

where *t* = days since pandemic has started in Italy

*h* = magnitude scale

*µ* = peak of daily cases (scale)

*σ* = standard deviation (shape)

So, given

- the parameter vector *θ* = (*h, µ, σ*), where *θ ∈* Θ
- the total active cases *x*_*t*_ since the infection spread
- a generic bootstrap distribution 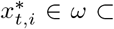 where *i {*1. *∈.N*} is the i-th bootstrap within N replicates
- 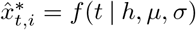 where 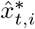 is the theoretical value

the objective is

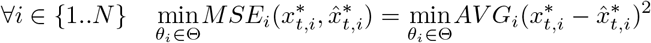

This is a non-linear unconstrained optimization problem which cannot be addressed using standard global optimization methods (Simplex, Branch and Bound or Branch and Cut algorithm), which are designed for Linear Programming LP (Murty (1983)) and Mixed-Integer Linear Programming MILP (Bénichou et al. (1971)), within the field of discrete combinatorial problems (Papadimitriou and Steiglitz (1998)).

Local search Simulated Annealing meta-heuristic to approximate global optimization can be used to solve unconstrained non-linear problems in a large space.

## 6. Simulated Annealing Optimization

Simulated annealing (SA), following Van Laarhoven and Aarts (1987), is a probabilistic technique for approximating the global optimum of a given function. Specifically, it is a metaheuristic used to approximate global optimization in a large search space for an optimization problem.

The name and inspiration come from annealing in metallurgy, a technique involving heating and controlled cooling of a material to increase the size of its crystals and reduce their defects. Both are attributes of the material that depend on its thermodynamic free energy. Heating and cooling the material affects both the temperature and the thermodynamic free energy. Simulated annealing can be used to approximate the global minimum for a function with many variables. In 1983, this approach was used by Kirkpatrick et al. (1983) for a solution of the traveling salesman problem. They also proposed its current name, simulated annealing.

This notion of slow cooling implemented in the simulated annealing algorithm is interpreted as a slow decrease in the probability of accepting worse solutions as the solution space is explored. Accepting worse solutions is a fundamental property of meta-heuristics because it allows for a more extensive search for the global optimal solution.

In general, simulated annealing algorithms work as below explained. The temperature progressively decreases from an initial positive value to zero. At each time step, the algorithm randomly selects some neighbor state *s*^*\**^ of the current state *s*, measures its energy (in this case the 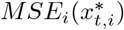 on the bootstrap distribution) and decides between moving the system to the state *s*^*\**^ or staying in state *s* according to the temperature-dependent probabilities of selecting better or worse solutions, which during the search respectively remain at 1 (or positive) and decrease towards zero.

### 6.1 Simulated annealing on bootstrap pseudocode

The following pseudocode presents the simulated annealing heuristic applied to bootstrap replicates. For each bootstrap it starts from a state *s*_0_ and continues searching solutions until temperature decay reaches a low temperature. In the process, the call Neighbour(*s, ϕ*) should generate a randomly chosen neighbour of a given state *s*; the call Random *U* (0, 1) should pick and return a value in the range [0, 1], uniformly at random. The annealing schedule is defined by the temperature decay based on the fixed cooling rate *ρ*.

- Let current temperature *T* = *t*_0_
- Set Cooling rate *ρ*
- For each bootstrap series *i* in {1, …, *N*}

− Let current solution *s* = *s*_0_
− Loop while temperature *T >* 1
* Pick a random neighbor, *s*_*new*_ *←* Neighbor(*s, ϕ*), where *ϕ* is the radius around *s*

* if *Prob*(*E*(*s*), *E*(*s*_*new*_) | *T, k*_*B*_) *≥* Random *U* (0, 1): *s* ← *s_new_*
* *T* ← *T* * (1 − *ρ*)
− Output the final state *s* on *i*-th bootstrap

where *Prob*(*E*(*s*), *E*(*s*_*new*_) | *T, k*_*B*_) is the acceptance probability at each iteration given temperature *T* and Boltzmann constant (see Aarts and Korst (1988)) *k*_*B*_

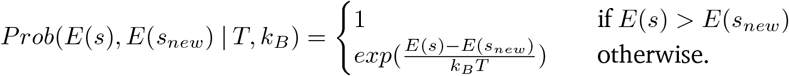

### 7. Empirical evidences

In order to improve local search speed the parameter space Θ can be bounded to Θ′ *⊂* Θ removing useless tails. No information is lost if parameters space is reduced to

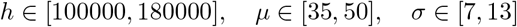

SA parameters has been recursively tuned and the procedure improved using an initial temperature *T*_0_ = 10000, a cooling rate *ρ* = 0.0006, Boltzmann constant *k*_*B*_ = 100 and radius *ϕ* = 0.3(*θ*_*max*_ *− θ*_*min*)_.

Optimization procedure applied to 500 Bootstraps, derived from active case in Italy from February 19^*th*^ to March 27^*th*^, shows with *Avg*(*h*) = 122178, *Avg*(*µ*) = 36.7, *Avg*(*σ*) = 10.8.

As an approximation, confidence bounds from normal distibution are derived for each of the parameters, such that with 0.01 significance level

**Fig. 2.**
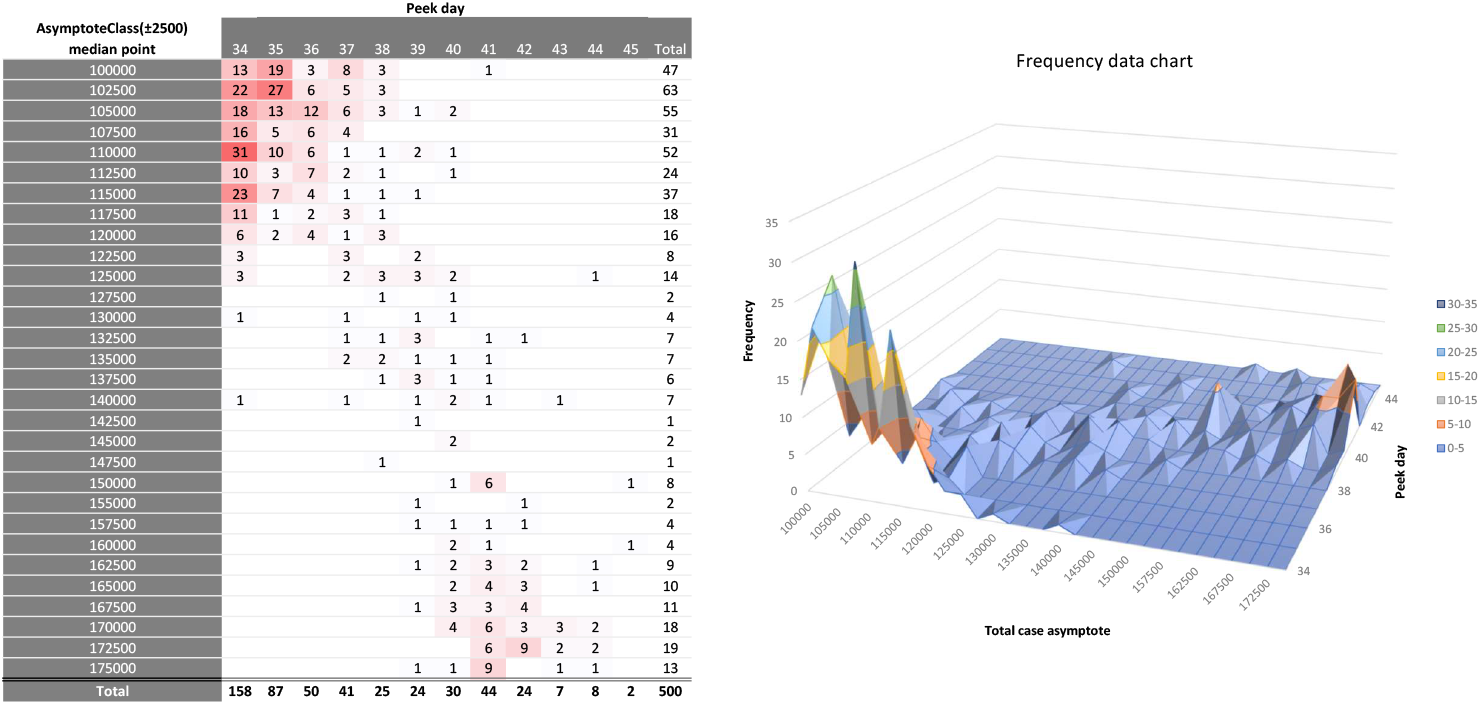
COVID19 bootstrap [AsymptoteClass,Peekday] frequency

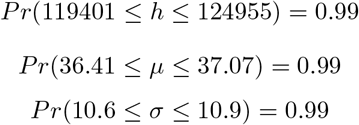

*µ* confidence interval points out a peak day of daily cases between March 25^*th*^ and 26^*th*^, while *h* magnitude parameter shows an asymptote of totale active cases curve between 120000 and 125000. The new cases curve has an asymptotic behavior, so cutting tail beyond a 0.1 cut-off for new infections, the pandemic time window is hypothetically over after May 16^*th*^.

This behavior is clearly described in the below charts built considering Gaussian and Cumulated Gaussian around 99% confidence lower and upper bound for each parameter.

**Fig. 3.**
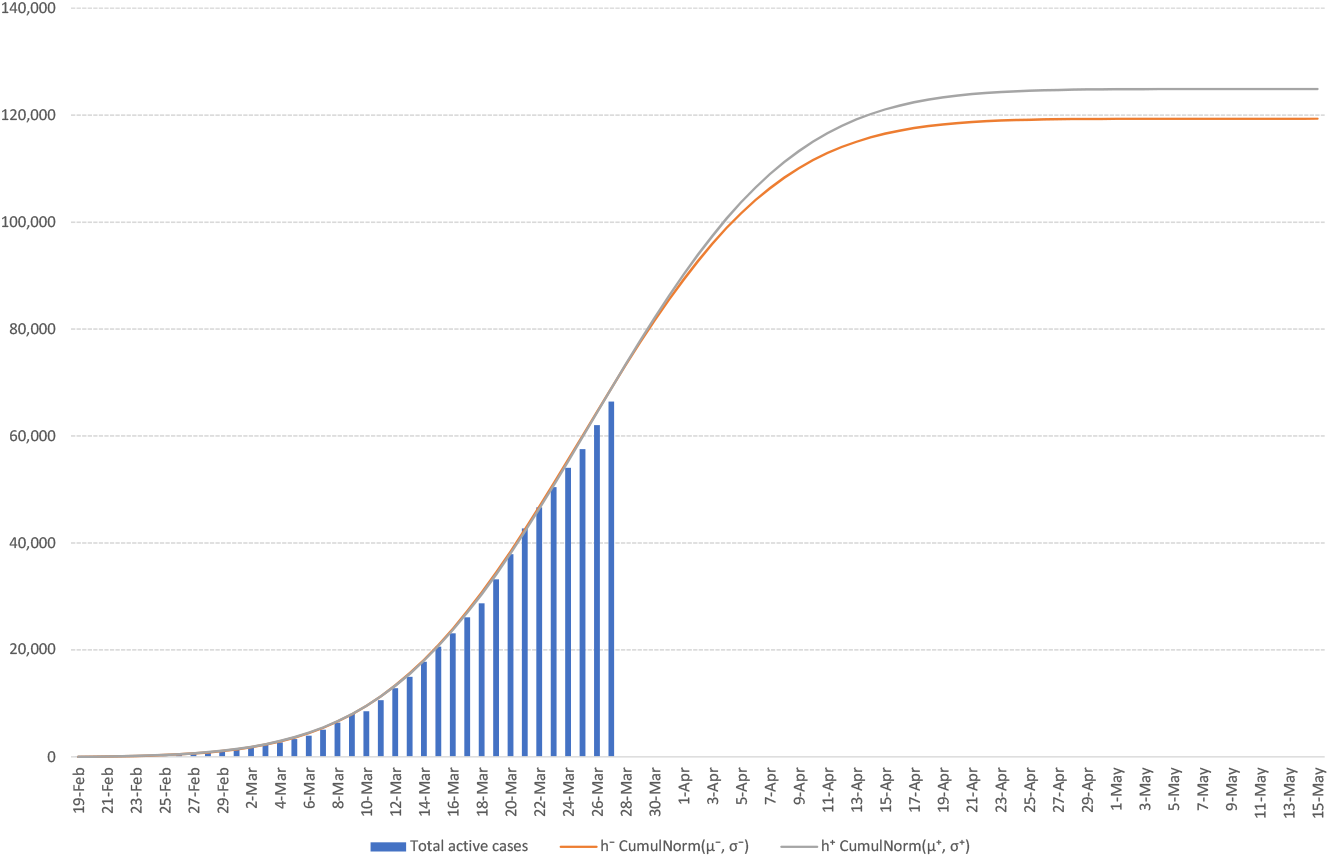
COVID19 Active cases fitting with Cumulated Gaussian

**Fig. 4.**
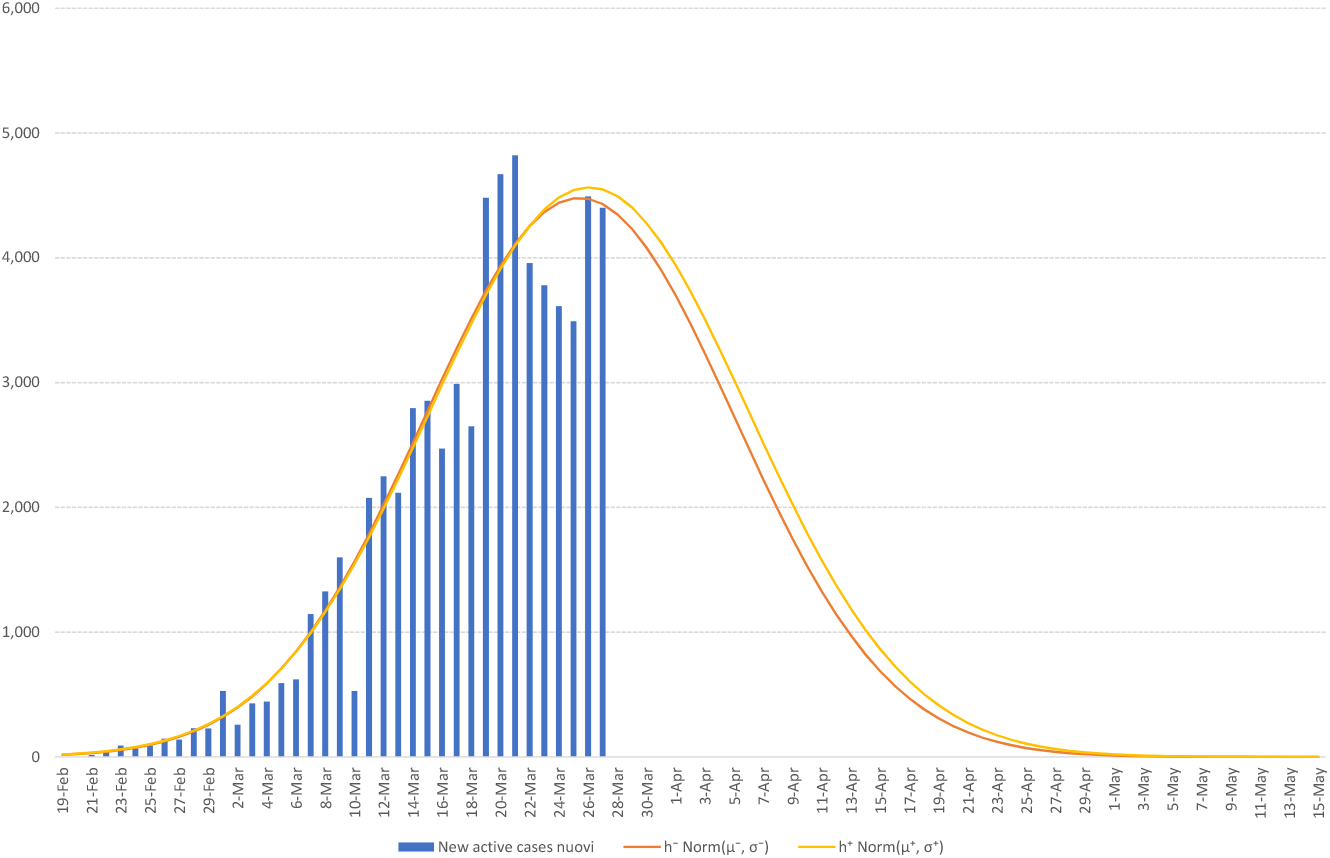
COVID19 New active cases fitting with Gaussian

## 8. Further developments

The SA optimization for fitting bootstraps derived from real data is applicable to any kind of distribution known in literature and empirical distributions as well.

This kind of research highlights a great potential if the aforementioned procedure is enhanced with the automatic choice of known distributions *ξ*_*r*_ or empirical ones *ξ*_*e*_ where (*ξ*_*r*_, *ξ*_*e*_) are in a predefined distribution space Ξ. More in detail, the algorithm could include a pre-processing light SA optimization (with an higher cooling rate *ρ* to cut down the number of SA iterations) to reduce the distribution space Ξ and the parameter space Θ_*ξ*_ for each distribution *ξ ∈* Ξ and boost the optimization search performed by the main SA process.

## 9. Disclaimer

The views and opinions expressed in this article are those of the authors and do not necessarily reflect the official policy or position of the Italian National Institute of Statistics or any other Entity.

## Data Availability

All the data are freely available at the web address
www.iss.it

